# Safety and immunogenicity of 23-valent pneumococcal polysaccharide vaccine (PPV23) in Chinese children, adults and elderly: a phase 4, randomized, double-blind, active-controlled clinical trial

**DOI:** 10.1101/2025.01.01.25319861

**Authors:** Xiaoyu Liu, Gang Shi, Wanqi Yang, Yuanyuan Dong, Xianying Ye, Juxiang Zhang, Xinyi Yang, Dan Song, Yuehong Ma, Zeng Wang, Hong Li, Dan Yu, Weijun Hu

## Abstract

**Objectives:** A randomized, double-blind, active-controlled noninferiority phase 4 clinical trial was conducted to evaluate the immunogenicity and safety of a 23-valent pneumococcal polysaccharide vaccine (PPV23).

**Methods:** Pneumococcal vaccine-naïve participants aged ≥2 years were randomly assigned in a 2:1 ratio to receive a single dose of either the treatment vaccine (n=1199) or a comparator vaccine (n=600). We evaluated the immunogenicity before and 30 days post-vaccination, by measuring serum IgG serotype-specific pneumococcal antibodies to 23 serotypes contained in the vaccines via an enzyme-linked immunosorbent assay. The primary outcome was seroconversion (two-fold increase) of serum IgG serotype-specific antibodies at days 30 compared with baseline.

**Results:** One month after the administration of PPV23, seroconversion rates for each of the 23 serotypes ranged from 59.22% to 95.67% in the treatment group, and in the control group from 59.66% to 94.07%. The lower bound of the 95% confidence interval (95%CI) of the rate differences for the 23 serotypes were all larger than −10%. Moreover, 12 serotypes (6B, 23F, 1, 2, 4, 8, 9N, 9V, 11A, 15B, 17F and 18C) had a lower bound of 95%CI for rate difference larger than 0. In total, 236 (19.68%) participants in the treatment group and 118 (19.67%) in the control group reported adverse reactions within 30 days poste-vaccination. No significant differences in incidence of adverse reactions were found between the two comparison groups.

**Conclusions:** The PPV23 vaccine administered among individual aged ≥2 years was safe, well tolerated and immunogenic, eliciting immune response either comparable to or higher than control vaccine.

## INTRODUCTION

Pneumococcal disease caused by Streptococcus pneumoniae (*S. pneumoniae,* or pneumococcus) is a major cause of morbidity and mortality worldwide, resulting in 1.19 million deaths of lower respiratory infection in 2016 [1]. The disease disproportionately affected young children and the elderly aged 65+ years, with extensive time, place and population variations [1]. *S. pneumoniae* usually colonizes the upper respiratory tract of healthy people, with approximately 5%-10% adult and 20%-60% children carry the bacterium in upper respiratory tract [2]. In certain susceptible populations (e.g., people with underlying chronic conditions, the elderly aged >65 years and infants) [3], it can spread to multiple organ systems and cause invasive pneumococcal disease (IPD) (bacteremia, meningitis, osteomyelitis, and septic arthritis) or noninvasive diseases (pneumonia without bacteremia, sinusitis, and otitis media) [4, 5]. There are at least 100 serotypes of *S. pneumoniae* [6], and the 24 serotypes contained in pneumococcal vaccines caused most of IPD diseases in children and adults (approximately 60%∼75%) [7, 8]. In China, systematic review found that IPD in children and adults were mainly caused by 6 serotypes, i.e. 3, 6B, 14, 19A, 19F and 23F [9–11]. Evaluating and monitoring the protective effect of pneumococcal vaccines routinely used in the population against different circulating serotypes after its licensure is an important measure and is essential for optimizing vaccine usage and for advocating future immunization practice and policies.

Pneumococcal vaccines are effective for the control and prevention of pneumococcal diseases [12, 13]. Currently, two types of pneumococcal vaccine are available in China, i.e., 13-valent polysaccharide conjugate vaccine (PCV13) for use in children aged 6 weeks to five years, and 23-valent pneumococcal polysaccharide vaccine (PPV23) for use in high-risk populations aged 2 years and older [14]. However, owing to the lack of domestic evidence on disease burden, health economic data or perhaps concerns of limited vaccine supply capacity, no pneumococcal vaccines were included in China’s national immunization program, but are administered on a private purchase basis and vaccine uptake remains suboptimal (3%-9%) [15, 16].

To increase domestic access to the pneumococcal vaccine, Sinovac developed the preservative-free PPV23, which was first approved by Chinese authority in 2020 for use in individuals aged 2 years and older, especially in high-risk populations (i.e., the elderly people, immunocompetent individual with underlying medical conditions, immunocompromised individual, functional or anatomic asplenia, individual with AIDS/HIV, cerebrospinal fluid leak, and other high-risk groups). The pre-marketing clinical data showed that the vaccine is safe, well tolerated and effective in preventing pneumococcal disease [17]. Post-marketing surveillance also did not indicate any unanticipated safety signals. We conducted this phase 4 trial to assess the immunogenicity, tolerability, and safety of Sinovac PPV23 compared to one domestic available PPV23 vaccine Pneumovax® (Merck & Co., Inc., Kenilworth, NJ, USA) in children, adults and the elderly population in China. The trial was conducted in response to a request from the National Medical Products Administration (NMPA) of China and was registered with ClinicalTrials.gov (NCT05477693).

## MATERIAL & METHODS

### Study design and participants

This randomized, double-blind, active-controlled noninferiority phase clinical trial evaluated the immunogenicity and safety of a 23-valent pneumococcal polysaccharide vaccine (PPV23) in Chinese population aged ≥2 years. The study was conducted in Linwei district, Shaanxi province, China between September 28^th^, 2022 and March 5^th^, 2023. Participants were randomly assigned in a 2:1 ratio to receive either the treatment vaccine or a domestic commercially available comparator vaccine. Written informed consent was obtained from each participant/participant’s guardian before enrolment. The trial was approved by the Ethics Committee of Shaanxi Center for Disease Control and Prevention (reference no., 2022-001-02) and was conducted in compliance with Good Clinical Practices and the ethical principles of the Declaration of Helsinki. Written informed consent was obtained from participants or children’s legal guardians prior to enrollment; adolescents aged 9–17 years also signed a written assent.

Inclusion criteria included being aged ≥2 years, in stable healthy status. Main exclusion criteria included previous history of laboratory-confirmed pneumococcal disease; receipt of a licensed or investigational pneumococcal vaccine; known history of severe allergies or reaction to vaccines or any component of vaccine; having severe neurological disease or psychosis; acute illness or axillary temperature of more than 37.0 °C; receipt of immunosuppressive therapy within previous 6 months before enrolment, receipt of blood or blood products within previous 3 months before enrolment; receipt of any live attenuated vaccine within 14 days, or any inactivated vaccine within 7 days. Women were excluded if they were currently pregnant, lactating, or expected to be pregnant during the study. The full eligibility criteria can be found in ClinicalTrials.gov (NCT05477693).

### Randomization and masking

After enrollment, eligible participants within each of the three age strata (i.e., 2-17 years, 18-59 years, and 60+ years) were randomly allocated to receive the treatment vaccine and the control vaccine in a 2:1 ratio. Specifically, an unmasked statistician generated the randomization list via a stratified permuted block randomization in SAS 9.4 (SAS Institute Inc., Cary, NC, USA). The unmasked study staff labeled the container of each treatment and control vaccine with a code from the randomization list. The container of treatment and control vaccine were identical in appearance, except the code number. Each participant was assigned a sequential number according to their sequence of enrolment and allocated the vaccine with the same code number on container. During the trial, the participant, investigator assessing safety and immunogenicity outcomes, and the sponsor were kept masked.

### Procedures

The treatment vaccine is a preservative-free 23-valent PPV (PPV23, Sinovac Biotech Co., Ltd., Beijing, China), a sterile liquid suspension of highly purified capsular polysaccharide antigens of S. pneumoniae serotypes 1, 2, 3, 4, 5, 6B, 7F, 8, 9N, 9V, 10A, 11A, 12F, 14, 15B, 17F, 18C, 19A, 19F, 20, 22F, 23F, and 33F. The treatment vaccine contains a mixture of 25 μg antigens for each serotype and was packaged in 0.5 ml prefilled syringe. The control vaccine was another domestic commercially available vaccine, Pneumovax® (Merck & Co., Inc., Kenilworth, NJ, USA). The control vaccine contains the same serotypes and amounts of polysaccharide antigens as the treatment vaccine and 0.25% phenol as a preservative. The treatment and control vaccines were administered once via intramuscular injection in the deltoid muscle of the upper arm in a volume of 0.5 mL.

After vaccination, all participants were monitored for immediate adverse events (AEs) for 30 minutes at the study site. AEs, including predefined symptoms (solicited AEs) and unspecified symptoms (unsolicited AEs) were collected on diary cards. Solicited AEs, including local AEs (e.g., injection site pain, induration, swelling, erythema, rash, and pruritus) and systemic AEs (e.g., fever, acute hypersensitive reaction, cough, myalgia, arthralgia, headache, and fatigue), were recorded within 7 days following vaccination, while unsolicited AEs were recorded within 30 days after vaccination. All participants (or guardians) were required to fill out diary card and spontaneously report AEs that occurred within 30 days after vaccination. Study investigators conducted a face-to-face interview on Day 7 to ensure completeness and accuracy of the safety data. Any serious adverse events (SAEs) were reported up to 30 days since enrollment. The study investigators decided relevance of the event with vaccination. Adverse events were graded according to the guideline provided by the National Medical Product Administration [18].

Blood samples were collected for immunogenicity analysis before injection (day 0) and at day 30 after vaccination. Sera were separated immediately and stored at −20 °C until analysis. Serum IgG serotype-specific pneumococcal antibodies to 23 serotypes contained in the vaccines were tested by enzyme-linked immunosorbent assay (ELISA) at the Chinese National Institute for Food and Drug Control, Beijing, China, as described previously [19].

### Outcomes

The primary immunogenicity outcomes were seroconversion, defined as the proportion of study participants who achieved at least two-fold increase of serum IgG serotype-specific pneumococcal antibodies at days 30 compared with baseline. The secondary immunogenicity outcomes included geometric mean concentration (GMC) and geometric mean fold increase (GMI) of log-transformed IgG antibodies to the 23 serotypes contained in the vaccines at day 30 post-vaccination.

The safety outcomes were based on incidence of solicited local AEs and systemic AEs, unsolicited AEs and SAEs, including proportion of participants reporting solicited local reactions within 7 days post-vaccination, solicited systemic reactions within 7 days post-vaccination, unsolicited AEs within 30 days post-vaccination, and SAEs within 30 days post-vaccination.

### Statistical analysis

The sample size of 1,800 was determined based on the assumed seroconversion rate of 65% in the control vaccine group for IgG antibodies to each of the six serotypes (3, 6B, 14, 19A, 19F, and 23F). Assuming a non-inferiority margin of −10%, a treatment to control ratio of 2:1 and a one-sided significance level of 0.025, we estimated that a minimum size of 956 participants in the treatment group and 478 in the control group would be required to achieve the overall power of 80% (power for each serotype=96.67%, calculated with Bonferroni method). Allowing a dropout rate to 20%, the final sample was 1200 in the treatment group and 600 in the control group.

We compared log-transformed GMI relative to baseline and GMC at day 30 with student’s t-test, while seroconversion rate at day 30 with Chi-square test or Fisher’s exact test. We calculated the rate differences of seroconversion between the treatment and control group and its 95% confidence interval (95% CI) using unstratified analysis of Miettinen-Nurminen method, and non-inferior was concluded if the lower bound of the 95% CI of rate differences for each serotype in the primary outcome were larger than −10%. Adverse events were summarized descriptively as frequencies and percentages by type of event and severity. Subgroup analyses were conducted, stratifying analysis by age groups (i.e., 2-17 years, 18-59 years, and 60+ years). A two-sided P-value of <0.05 was considered statistically significant. We conducted all analyses in SAS (version 9.4, SAS Institute Inc., Cary, NC, USA).

## RESULTS

### Study population

In total, 2039 participants were screened for eligibility, of whom 1800 were enrolled and randomly allocated. One participant did not receive the allocated vaccine after randomization. 1199 received treatment vaccine and 600 received control vaccine, and they were included in the safety analysis population (n=1799). 1177 participants in the treatment group and 590 participants in the control group were included in the immunogenicity analysis population (n=1767). 32 participants were excluded because 22 had no blood drew post-vaccination, 3 participants had blood drew but out of the visit window, 6 participants met the exclusion criteria, and 1 participant in control group received the wrong vaccine (Figure 1). Baseline characteristics, i.e., age, sex, ethnic, BMI and underlying medical conditions, were similar between the treatment and control group (Table 1). The baseline serum IgG antibody GMCs against each of the 23 pneumococcal serotypes, ranging from 0.32 to 6.95 in the treatment group and 0.34 to 6.93 in the control group, were not significantly different at baseline (all p>0.05) (Table 2), suggesting balance between the two comparison groups before vaccination.

**Figure 1.**
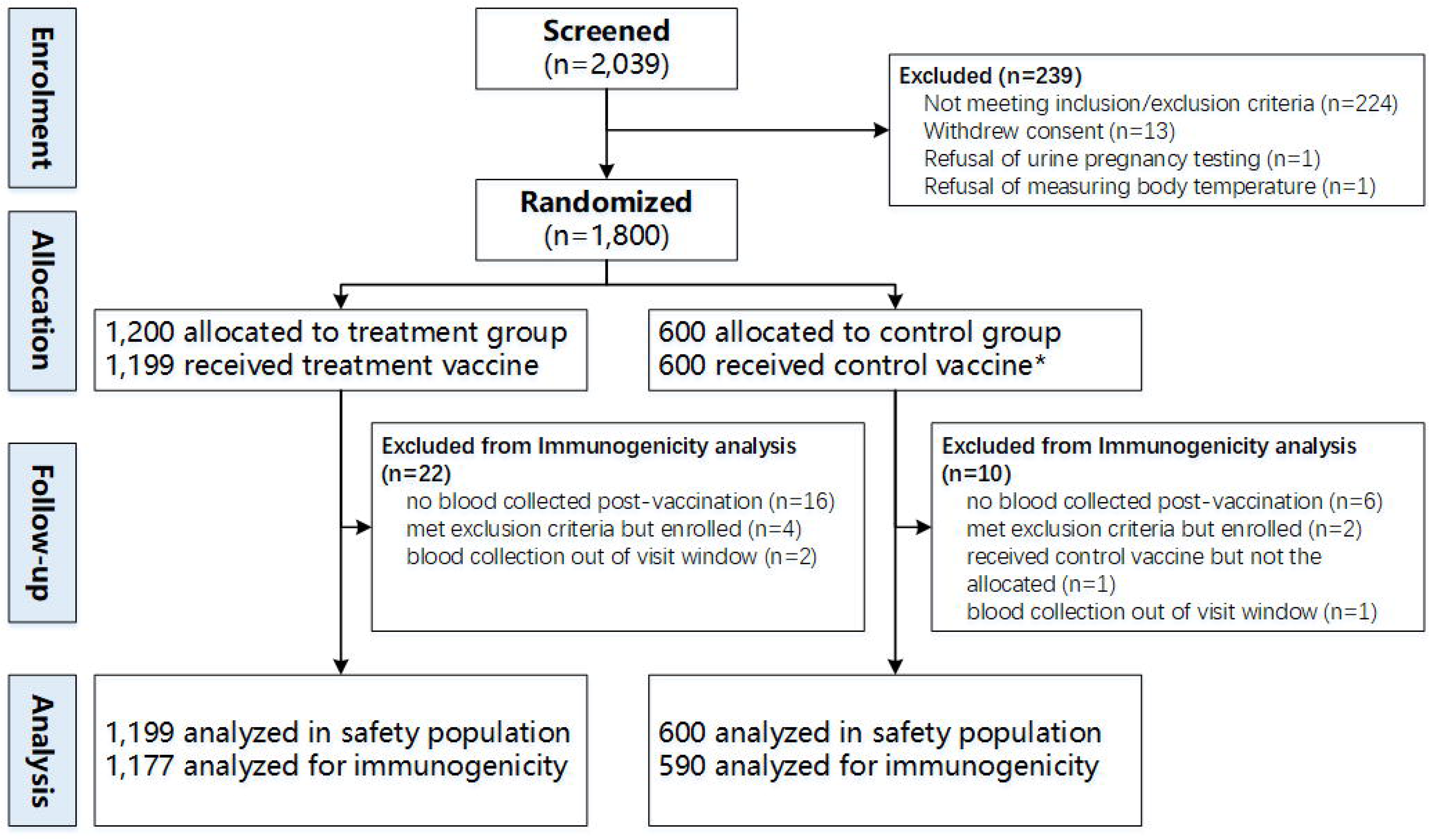
Trial profile. *One participant in control group received the other control vaccine, not the randomly allocated one.

**Table 1.** Participant population demographic characteristics.

| Characteristics | Safety population, no. (%) |  |
| --- | --- | --- |
|  | Treatment | Control |
| No. of participants | 1199 | 600 |
| Mean age in years (SD) | 33.5(25.56) | 33.7(25.50) |
| Age group |  |  |
| 2-17 years | 599(49.96) | 300(50.00) |
| 18-59 years | 240(20.02) | 121(20.17) |
| 60+ years | 360(30.03) | 179(29.83) |
| Sex |  |  |
| Male | 582(48.54) | 297(49.50) |
| Female | 617(51.46) | 303(50.50) |
| Ethnicity |  |  |
| Han nationality | 1197(99.83) | 599(99.83) |
| Others | 2(0.17) | 1(0.17) |
| BMI, kg/m<sup>2</sup> (SD) | 22.11(4.86) | 22.08(4.66) |
| Underlying medical conditions | 125(10.42) | 67(11.17) |

Note: SD=standard deviation; BMI=body mass index.

**Table 2.** Serological Response of study participants to the 23-valent Pneumococcal Polysaccharide Vaccines.

| Serotypes | GMC (95%CI) at baseline |  | p-value | GMC (95%CI) post-vaccination |  | GMC ratio<br>(95%CI)<br>post-vaccination | p-value | GMI (95%CI) post-vaccination |  | p-value |
| --- | --- | --- | --- | --- | --- | --- | --- | --- | --- | --- |
|  | Treatment<br>(n=1177) | Control<br>(n=590) |  | Treatment<br>(n=1177) | Control<br>(n=590) |  |  | Treatment<br>(n=1177) | Control<br>(n=590) |  |
| 3 | 0.35(0.32, 0.38) | 0.37(0.34, 0.42) | 0.2858 | 1.06(1.01, 1.12) | 1.21(1.13, 1.29) | 0.88(0.81,0.96) | 0.0035 | 3.04(2.85, 3.25) | 3.22(2.93, 3.54) | 0.3387 |
| 6B | 2.04(1.94, 2.14) | 2.04(1.91, 2.19) | 0.9505 | 6.92(6.48, 7.39) | 6.45(5.88, 7.08) | 1.07(0.96,1.20) | 0.227 | 3.39(3.22, 3.58) | 3.15(2.93, 3.40) | 0.1182 |
| 14 | 6.95(6.64, 7.27) | 6.93(6.50, 7.38) | 0.9435 | 33.07(31.01, 35.28) | 30.64(27.97, 33.56) | 1.08(0.97,1.21) | 0.1795 | 4.76(4.47, 5.07) | 4.42(4.04, 4.84) | 0.1873 |
| 19F | 3.36(3.22, 3.51) | 3.41(3.21, 3.63) | 0.6912 | 14.23(13.43, 15.09) | 12.95(11.92, 14.06) | 1.1(0.99,1.22) | 0.0664 | 4.24(4.02, 4.46) | 3.8(3.53, 4.09) | 0.0171 |
| 19A | 3.71(3.54, 3.90) | 3.79(3.54, 4.06) | 0.624 | 12.98(12.26, 13.75) | 12.31(11.34, 13.35) | 1.06(0.96,1.17) | 0.2909 | 3.5(3.31, 3.69) | 3.25(3.01, 3.51) | 0.1198 |
| 23F | 1.51(1.44, 1.58) | 1.46(1.36, 1.56) | 0.4271 | 6.43(6.05, 6.84) | 5.35(4.91, 5.84) | 1.2(1.08,1.34) | 0.0008 | 4.27(4.04, 4.50) | 3.67(3.40, 3.97) | 0.0017 |
| 1 | 0.75(0.70, 0.80) | 0.79(0.71, 0.86) | 0.4281 | 6.13(5.76, 6.52) | 5.91(5.41, 6.45) | 1.04(0.93,1.16) | 0.4993 | 8.17(7.64, 8.74) | 7.52(6.84, 8.26) | 0.1564 |
| 2 | 3.42(3.27, 3.58) | 3.54(3.31, 3.78) | 0.4186 | 25.06(23.81, 26.38) | 21.13(19.65, 22.71) | 1.19(1.09,1.30) | 0.0002 | 7.32(6.95, 7.71) | 5.97(5.55, 6.43) | <0.0001 |
| 4 | 0.43(0.40, 0.47) | 0.42(0.38, 0.47) | 0.7733 | 3.65(3.47, 3.84) | 2.67(2.48, 2.87) | 1.37(1.25,1.49) | <0.0001 | 8.46(7.78, 9.20) | 6.32(5.61, 7.11) | <0.0001 |
| 5 | 0.61(0.58, 0.65) | 0.63(0.58, 0.68) | 0.6485 | 4.46(4.22, 4.72) | 4.11(3.79, 4.45) | 1.09(0.99,1.20) | 0.0962 | 7.29(6.86, 7.75) | 6.56(6.02, 7.15) | 0.0494 |
| 7F | 1.17(1.11, 1.25) | 1.21(1.11, 1.32) | 0.5463 | 7.38(6.94, 7.83) | 6.77(6.22, 7.38) | 1.09(0.98,1.21) | 0.11 | 6.28(5.91, 6.67) | 5.59(5.13, 6.08) | 0.027 |
| 8 | 2.15(2.04, 2.27) | 2.16(2.00, 2.33) | 0.9238 | 18.61(17.72, 19.54) | 14.8(13.81, 15.86) | 1.26(1.15,1.37) | <0.0001 | 8.66(8.14, 9.22) | 6.86(6.28, 7.49) | <0.0001 |
| 9N | 1.14(1.05, 1.25) | 1.19(1.05, 1.35) | 0.6068 | 12.02(11.35, 12.73) | 10.16(9.37, 11.02) | 1.18(1.07,1.31) | 0.0009 | 10.5(9.67, 11.41) | 8.53(7.59, 9.58) | 0.0043 |
| 9V | 1.49(1.40, 1.60) | 1.54(1.39, 1.69) | 0.6526 | 7.26(6.85, 7.69) | 6.39(5.88, 6.94) | 1.14(1.03,1.26) | 0.013 | 4.86(4.59, 5.14) | 4.16(3.84, 4.50) | 0.0018 |
| 10A | 2.89(2.77, 3.02) | 2.89(2.72, 3.07) | 0.9676 | 10.97(10.27, 11.71) | 9.86(8.98, 10.81) | 1.11(0.99,1.25) | 0.0653 | 3.79(3.60, 3.99) | 3.41(3.17, 3.67) | 0.0197 |
| 11A | 1.9(1.77, 2.03) | 2.04(1.85, 2.25) | 0.2446 | 6.67(6.36, 6.99) | 5.66(5.29, 6.05) | 1.18(1.09,1.28) | <0.0001 | 3.51(3.32, 3.72) | 2.78(2.56, 3.01) | <0.0001 |
| 12F | 0.32(0.30, 0.34) | 0.34(0.31, 0.37) | 0.2162 | 2.27(2.14, 2.41) | 2.25(2.07, 2.45) | 1.01(0.91,1.12) | 0.8544 | 7.18(6.73, 7.67) | 6.63(6.04, 7.27) | 0.1647 |
| 15B | 3.15(2.97, 3.34) | 3.14(2.89, 3.41) | 0.9561 | 20.68(19.35, 22.09) | 17.06(15.54, 18.73) | 1.21(1.08,1.36) | 0.001 | 6.56(6.19, 6.96) | 5.43(5.00, 5.90) | 0.0003 |
| 17F | 1.25(1.17, 1.33) | 1.31(1.20, 1.43) | 0.3613 | 8.45(7.99, 8.93) | 7.95(7.35, 8.60) | 1.06(0.96,1.17) | 0.2192 | 6.77(6.38, 7.19) | 6.07(5.58, 6.60) | 0.0369 |
| 18C | 1.26(1.17, 1.35) | 1.37(1.24, 1.52) | 0.1741 | 7.49(7.10, 7.90) | 6.7(6.20, 7.23) | 1.12(1.02,1.23) | 0.0189 | 5.96(5.59, 6.36) | 4.88(4.45, 5.35) | 0.0005 |
| 20 | 2.94(2.78, 3.11) | 2.99(2.77, 3.23) | 0.7141 | 10(9.45, 10.58) | 10.43(9.63, 11.29) | 0.96(0.87,1.06) | 0.404 | 3.4(3.23, 3.58) | 3.48(3.24, 3.74) | 0.5965 |
| 22F | 2.23(2.15, 2.32) | 2.24(2.12, 2.37) | 0.9242 | 8.48(8.08, 8.90) | 8.23(7.68, 8.81) | 1.03(0.95,1.12) | 0.4751 | 3.8(3.61, 4.00) | 3.67(3.42, 3.94) | 0.4484 |
| 33F | 1.11(1.01, 1.22) | 1.2(1.06, 1.37) | 0.3058 | 14.08(13.18, 15.03) | 12.69(11.57, 13.92) | 1.11(0.99,1.24) | 0.073 | 12.72(11.72,13.8) | 10.53(9.38, 11.82) | 0.0092 |

Note: GMC= geometric mean concentration; GMI= geometric mean fold increase; 95%CI= 95% confidence interval.

### Immunogenicity post vaccination

After vaccination, the IgG antibodies to the 23 *S. pneumoniae* serotypes contained in both vaccines were increased significantly compared with baseline (Table 2). The GMCs at days 30 post-vaccination ranged from 1.06 to 33.07 in the treatment group and 1.21 to 30.64 in the control group, increasing on average 3.04∼12.72 fold relative to baseline in the treatment group and 2.78∼10.53 fold in the control group, respectively. In both groups, serotype 14 had the highest GMCs among all the 23 serotypes assayed, while serotype 3 had the lowest. The GMCs ratio between treatment group and control group post-vaccination ranged from 0.88 to 1.37, with serotype 3 had the lowest (0.88, 95%CI=0.81,0.96) and serotype 4 had the highest (1.37, 95%CI=1.25,1.49). There were nine serotypes (2, 4, 8, 9N, 9V, 11A, 15B, 18C, and 23F) whose GMCs’ 95% CI had a lower bounder of larger than 1 (Table 2).

Regarding the seroconversion (≥2-fold increase at days 30 post-vaccination), the seroconversion rate for each of the 23 serotypes ranged from 59.22% to 95.67% in the treatment group, and in the control group from 59.66% to 94.07%, respectively. The lower bound of the 95% CI of seroconversion rate differences (treatment group minus control group) of IgG antibodies for all 23 serotypes were all larger than −10%. Moreover, we found that there were an additional 12 serotypes (1, 2, 4, 8, 6B, 9N, 9V, 11A, 15B, 17F, 18C, and 23F), which had a lower bound of 95%CI for rate difference larger than 0 (Table 3).

**Table 3.** Seroconversion rate of study participants inoculated with the 23-valent Pneumococcal Polysaccharide Vaccines.

| Serotypes | Treatment group (n=1177) |  | Control group (n=590) |  | rate difference (95% CI) | p-value |
| --- | --- | --- | --- | --- | --- | --- |
|  | no. | rate (95% CI) | no. | rate (95% CI) |  |  |
| 3 | 697 | 59.22(56.35,62.04) | 358 | 60.68(56.61,64.64) | -1.46(-6.26,3.41) | 0.5552 |
| 6B | 823 | 69.92(67.21,72.53) | 374 | 63.39(59.36,67.29) | 6.53(1.89,11.26) | 0.0056 |
| 14 | 911 | 77.40(74.90,79.76) | 443 | 75.08(71.39,78.53) | 2.32(-1.83,6.63) | 0.2781 |
| 19F | 933 | 79.27(76.84,81.55) | 456 | 77.29(73.69,80.61) | 1.98(-2.03,6.17) | 0.3382 |
| 19A | 843 | 71.62(68.95,74.18) | 402 | 68.14(64.21,71.88) | 3.49(-1.01,8.10) | 0.1297 |
| 23F | 931 | 79.10(76.66,81.39) | 431 | 73.05(69.28,76.59) | 6.05(1.86,10.39) | 0.0043 |
| 1 | 1096 | 93.12(91.52,94.50) | 533 | 90.34(87.66,92.60) | 2.78(0.12,5.74) | 0.0400 |
| 2 | 1104 | 93.80(92.26,95.11) | 525 | 88.98(86.17,91.39) | 4.81(2.08,7.87) | 0.0004 |
| 4 | 1046 | 88.87(86.93,90.61) | 482 | 81.69(78.33,84.73) | 7.18(3.69,10.91) | <0.0001 |
| 5 | 1098 | 93.29(91.70,94.65) | 546 | 92.54(90.12,94.53) | 0.75(-1.70,3.48) | 0.5613 |
| 7F | 1049 | 89.12(87.21,90.85) | 508 | 86.10(83.04,88.79) | 3.02(-0.17,6.48) | 0.0640 |
| 8 | 1099 | 93.37(91.80,94.73) | 523 | 88.64(85.80,91.09) | 4.73(1.94,7.83) | 0.0006 |
| 9N | 1077 | 91.50(89.76,93.03) | 521 | 88.31(85.43,90.79) | 3.2(0.28,6.40) | 0.0311 |
| 9V | 1023 | 86.92(84.85,88.79) | 475 | 80.51(77.08,83.63) | 6.41(2.78,10.26) | 0.0004 |
| 10A | 860 | 73.07(70.43,75.58) | 405 | 68.64(64.73,72.37) | 4.42(-0.04,9.00) | 0.0519 |
| 11A | 815 | 69.24(66.52,71.87) | 352 | 59.66(55.58,63.65) | 9.58(4.86,14.36) | <0.0001 |
| 12F | 1066 | 90.57(88.75,92.18) | 534 | 90.51(87.85,92.75) | 0.06(-2.73,3.11) | 0.9672 |
| 15B | 1043 | 88.62(86.66,90.37) | 496 | 84.07(80.86,86.93) | 4.55(1.20,8.15) | 0.0072 |
| 17F | 1081 | 91.84(90.13,93.34) | 525 | 88.98(86.17,91.39) | 2.86(0.01,6.00) | 0.0488 |
| 18C | 1034 | 87.85(85.85,89.66) | 483 | 81.86(78.51,84.89) | 5.99(2.47,9.74) | 0.0007 |
| 20 | 847 | 71.96(69.30,74.51) | 409 | 69.32(65.43,73.02) | 2.64(-1.82,7.22) | 0.2483 |
| 22F | 871 | 74.00(71.40,76.49) | 426 | 72.20(68.40,75.78) | 1.8(-2.53,6.27) | 0.4198 |
| 33F | 1126 | 95.67(94.34,96.76) | 555 | 94.07(91.85,95.83) | 1.6(-0.50,4.03) | 0.1407 |

### Subgroup analysis

Subgroup immunogenicity analysis by age groups showed that GMCs of IgG antibodies against each of 23 serotypes increased with age at baseline, with elderly aged 60+ years the highest, adults aged 18-59 years moderate, and children aged 2-17 years the lowest (*Supplementary Table 1-3*). Except for a few serotypes (i.e., Serotype 3 in children aged 2-17 years), the difference of seroconversion rate for most of the 23 serotypes in each age group achieved prespecified immunogenicity non-inferior criteria (Figure 2). Wider confidence intervals were observed for seroconversion rate differences in the separate analysis, because of reduced sample size and lowered statistical power.

**Figure 2.**
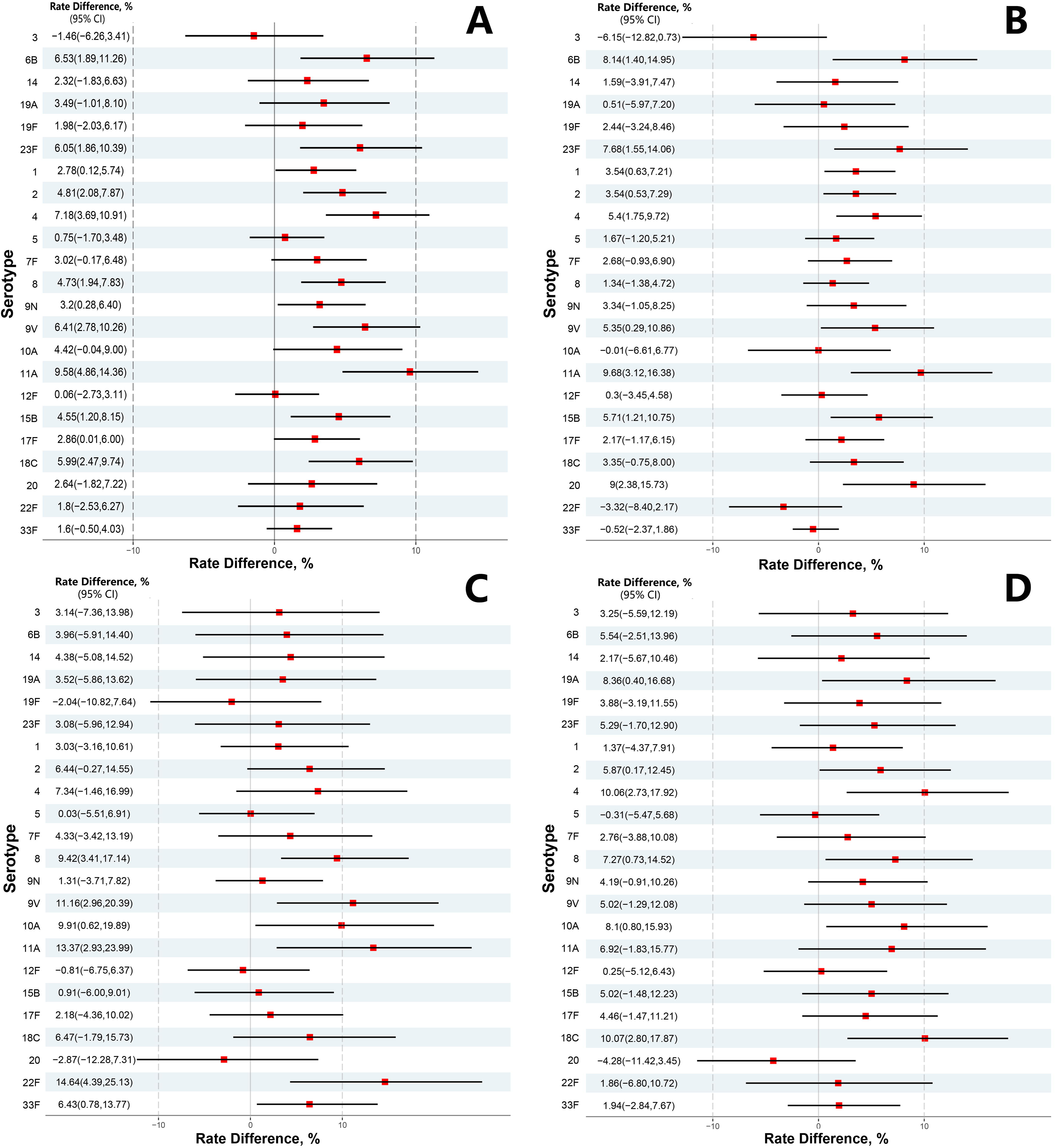
Seroconversion rate difference after vaccination. (A) all participants; (B) children aged 2∼17 years; (C) adults aged 18∼59 years; (D) elderly people aged 60+ years.

### Adverse reactions

In total, 236 (19.68%) participants in the treatment group and 118 (19.67%) in the control group reported adverse reactions within 30 days after vaccination. No significant differences in incidence of adverse reactions were found between the two comparison groups. The most common injection-site and systemic adverse reaction was vaccination site pain and fever. Most of adverse reactions were Grade 1 and Grade 2 events in severity and Grade 3 events were reported in 8 (0.67%) participants in the treatment group and 5 (0.83%) in the control group (p=0.7697). No deaths or grade 4 or above adverse reactions were reported during the study period. The most common reported Grade 3 adverse reaction was fever (Table 4). During the study period, 9 episodes of SAEs were reported by 6 participants, and all reported in the treatment group. Except one episodes of SAEs (urticaria with high fever occurred on days 20 after vaccination) were determined by the study investigator as related to the vaccination, the rest of SAEs were considered unrelated.

**Table 4.**
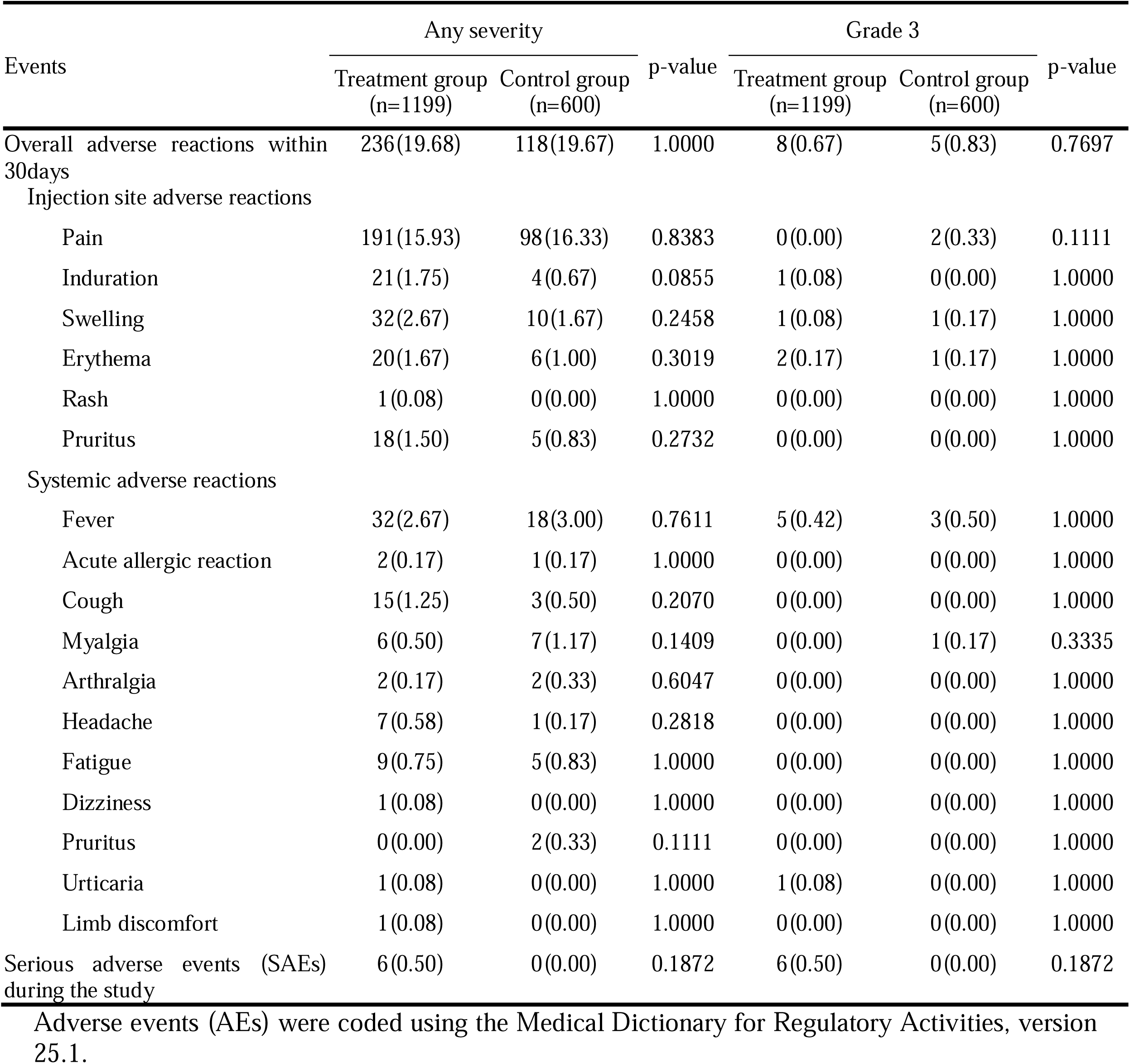
Summary of Adverse Reactions in the safety analysis population.

| Events | Any severity |  | p-value | Grade 3 |  | p-value |
| --- | --- | --- | --- | --- | --- | --- |
|  | Treatment group<br>(n=1199) | Control group<br>(n=600) |  | Treatment group<br>(n=1199) | Control group<br>(n=600) |  |
| Overall adverse reactions within 30days | 236(19.68) | 118(19.67) | 1.0000 | 8(0.67) | 5(0.83) | 0.7697 |
| Injection site adverse reactions |  |  |  |  |  |  |
| Pain | 191(15.93) | 98(16.33) | 0.8383 | 0(0.00) | 2(0.33) | 0.1111 |
| Induration | 21(1.75) | 4(0.67) | 0.0855 | 1(0.08) | 0(0.00) | 1.0000 |
| Swelling | 32(2.67) | 10(1.67) | 0.2458 | 1(0.08) | 1(0.17) | 1.0000 |
| Erythema | 20(1.67) | 6(1.00) | 0.3019 | 2(0.17) | 1(0.17) | 1.0000 |
| Rash | 1(0.08) | 0(0.00) | 1.0000 | 0(0.00) | 0(0.00) | 1.0000 |
| Pruritus | 18(1.50) | 5(0.83) | 0.2732 | 0(0.00) | 0(0.00) | 1.0000 |
| Systemic adverse reactions |  |  |  |  |  |  |
| Fever | 32(2.67) | 18(3.00) | 0.7611 | 5(0.42) | 3(0.50) | 1.0000 |
| Acute allergic reaction | 2(0.17) | 1(0.17) | 1.0000 | 0(0.00) | 0(0.00) | 1.0000 |
| Cough | 15(1.25) | 3(0.50) | 0.2070 | 0(0.00) | 0(0.00) | 1.0000 |
| Myalgia | 6(0.50) | 7(1.17) | 0.1409 | 0(0.00) | 1(0.17) | 0.3335 |
| Arthralgia | 2(0.17) | 2(0.33) | 0.6047 | 0(0.00) | 0(0.00) | 1.0000 |
| Headache | 7(0.58) | 1(0.17) | 0.2818 | 0(0.00) | 0(0.00) | 1.0000 |
| Fatigue | 9(0.75) | 5(0.83) | 1.0000 | 0(0.00) | 0(0.00) | 1.0000 |
| Dizziness | 1(0.08) | 0(0.00) | 1.0000 | 0(0.00) | 0(0.00) | 1.0000 |
| Pruritus | 0(0.00) | 2(0.33) | 0.1111 | 0(0.00) | 0(0.00) | 1.0000 |
| Urticaria | 1(0.08) | 0(0.00) | 1.0000 | 1(0.08) | 0(0.00) | 1.0000 |
| Limb discomfort | 1(0.08) | 0(0.00) | 1.0000 | 0(0.00) | 0(0.00) | 1.0000 |
| Serious adverse events (SAEs) during the study | 6(0.50) | 0(0.00) | 0.1872 | 6(0.50) | 0(0.00) | 0.1872 |

Adverse events (AEs) were coded using the Medical Dictionary for Regulatory Activities, version 25.1.

## DISCUSSION

The World Health Assembly endorsed the Immunization Agenda 2030 (IA2030) in August 2020, setting up seven strategic priorities to expand immunization delivery across the life course, reach high equitable immunization coverage, and build sustained supply and access to vaccines [20]. China has a substantial burden of pneumococcal disease [21, 22]. Conducting post-marketing study on the approved vaccines can secure safe and effective use of the product in the population, optimizing immunization practice and recommendations, as well as improving public awareness on equitable access to the vaccine. In this phase 4 trial, we evaluated the safety and immunogenicity of one PPV23 in a wide age group and large sample. Our findings showed that the Sinovac PPV23 vaccine is immunologically non-inferior to active control vaccine, with lower bound of the 95% CI for seroconversion rate differences to all the 23 serotypes achieved prespecified criteria (>-10%). The non-inferiority of the tested PPV23 vaccine to active control vaccine can be readily claimed. Moreover, there were 12 serotypes that had a lower bound of 95%CI larger than 0 for seroconversion rate differences, suggesting these antigens might elicit a superior IgG antibodies response.

Our immunogenicity results in this post-market study were consistent with one pivotal phase 3 clinical trial of the studied vaccine, in which the IgG antibody response was compared with another active control PPV23 vaccine (Chengdu Institute of Biological Products Co., Ltd.) [17]. The seroconversion rate (2-fold increase post-vaccination) of the treatment vaccine for 23 serotypes ranged from 49.71% to 90.96%, similar with this study (from 59.22% to 95.67%). In that study, all the 23 serotypes achieved immunogenicity non-inferior criteria for seroconversion rate, and 10 serotypes (1, 2, 6B, 8, 9N, 9V, 15B, 17F, 18C, and 23F) had superior antibodies response. These results and serotypes were repeatedly observed in the current study, suggesting a real effect was possessed by Sinovac PPV23. Interestingly, in the previous phase 3 trial did not meet the non-inferior criteria of immunogenicity for 10 serotypes (i.e., 3, 5, 7F, 8, 11A, 12F, 14, 15B, 19A and 19F) among children aged 2-17 years in subgroup analysis. The investigators reasoned that this is owing to a small sample size and insufficient statistical power in the children group in that trial. In our study with carefully calculated and enlarged sample size in children, 5 of the 6 above serotypes (except Serotype 3) achieved immunogenicity non-inferior with the control vaccine, eliminating our concerns over the efficacy and immunogenicity of the treatment vaccine in children in previous study. Indeed, Serotype 3 is the antigen component in the vaccine that induced the lowest IgG GMCs in both the treatment group (1.06, 95%CI=1.01-1.12) and control group (1.21, 95%CI=1.13-1.29).

The safety profile of the treatment vaccine was also similar with results in previous studies on PPV23 [17, 23, 24]. The most common local and systemic adverse reactions were pain and fever, and most of these events occurred within 7 days and were mild in severity (i.e., grade 1 and grade 2 events). Although the frequency of adverse reactions varied between studies, no significant differences were observed between active controls, suggesting a variation in study population, events definition might exist between studies. One serious adverse event of urticaria was determined by the investigator as related to the vaccination, which occurred 20 days after vaccination. World Health Organization guidance mentioned that severe allergic reactions (including urticaria and vascular edema, etc.) caused by immunization usually occurs within a few minutes after vaccination, rarely within 2 hours after vaccination, and delayed allergic reactions may occur within 48 hours[25]. Considering the longer interval from vaccination, accompanied with fever and elevated white blood cells count, this event was possibly related to infectious disease rather than vaccination.

Our study has several limitations. First, our observation period (30 days) was too short to evaluate immune persistence. The previous studies showed that PPV23 induced protection declining fast, with most studies having a duration of 2 to 5 years [12, 26, 27]. We will need to obtain the duration of protection in future. Second, we intentionally excluded participants with a prior history of any pneumococcal vaccine before enrolment. We were unable to evaluate the benefits of sequential immunization schedule with PCV followed by PPV23. Finally, the trial did not evaluate the protective effect of the test vaccine against clinical outcomes. The effectiveness of the vaccine in preventing against IPD, pneumonia or acute otitis media should be further assessed in future.

## CONCLUSIONS

In summary, the tested PPV23 vaccine, administered once among individual aged ≥2 years and who had no prior history of pneumococcal vaccines, was safe, well tolerated and immunogenic, eliciting immune response either comparable to or higher than the active control vaccine. The findings of this study support the use of the PPV23 in population aged ≥2 years.

### Abbreviations

AE: Adverse event; CDC: Center for Disease Control and Prevention; ELISA: Enzyme-Linked Immune-sorbent Assay; GMC: Geometric Mean Concentration; GMI: Geometric Mean Fold Increase; IPD: invasive pneumococcal disease; NMPA: National Medical Products Administration; PCV: polysaccharide conjugate vaccine; PPV: Pneumococcal polysaccharide vaccine; SAE: Serious adverse event; 95% CI: 95% confidence interval.

## Data Availability

All data produced in the present study are available upon reasonable request to the authors.

## Acknowledgements

We thank the subjects who participated in this study, their parents and guardians, and those who conducted the study, including study investigators, nurses, physicians, coordinators, and the clinical research associates and scientists at Sinovac. XL, GS, and WY contributed equally to this work. XL, DY, and WH designed the trial and study protocol. YD, XY (Xinyi Yang), and JZ contributed to the literature search. XL, YM, and ZW verified the data. XL, GS, and WY wrote the first draft manuscript. DY, WH, XY (Xinyi Yang), YDand ZW contributed to the data interpretation and revision of the manuscript. WY, and YM contributed to data analysis. XY (Xianying Ye) monitored the trial. DS and JZ were responsible for the site work including recruitment, follow-up, and data collection. GS and HL were responsible for the laboratory analysis. All the authors had full access to all the data in the study and had final responsibility for the decision to submit for publication.

## Patient and public involvement

Patients and/or the public were not involved in the design, or conduct, or reporting, or dissemination plans of this research.

## Patient consent for publication

Not required.

## Data sharing

De-identified individual participant-level data will be available upon written request to the corresponding author following publication. Requests must be accompanied by a detailed protocol and statistical analysis plan and will be reviewed for scientific validity. For those requestors whose proposals meet the research criteria, and for which an exception does not apply, data will be transferred to requestors via a secure portal. To gain access, data requestors must enter into a data sharing agreement with Sinovac.

## Ethics approval

This study was approved by the Ethics Committee of Shaanxi Center for Disease Control and Prevention (reference no., 2022-001-02).

## Supplementary Materials

This appendix has been provided by the authors to give readers additional information about their work.

Supplement to: Safety and immunogenicity of 23-valent pneumococcal polysaccharide vaccine (PPV23) in Chinese children, adults and elderly: a phase 4, randomized, double-blind, active-controlled clinical trial.

**Supplementary Table 1.**
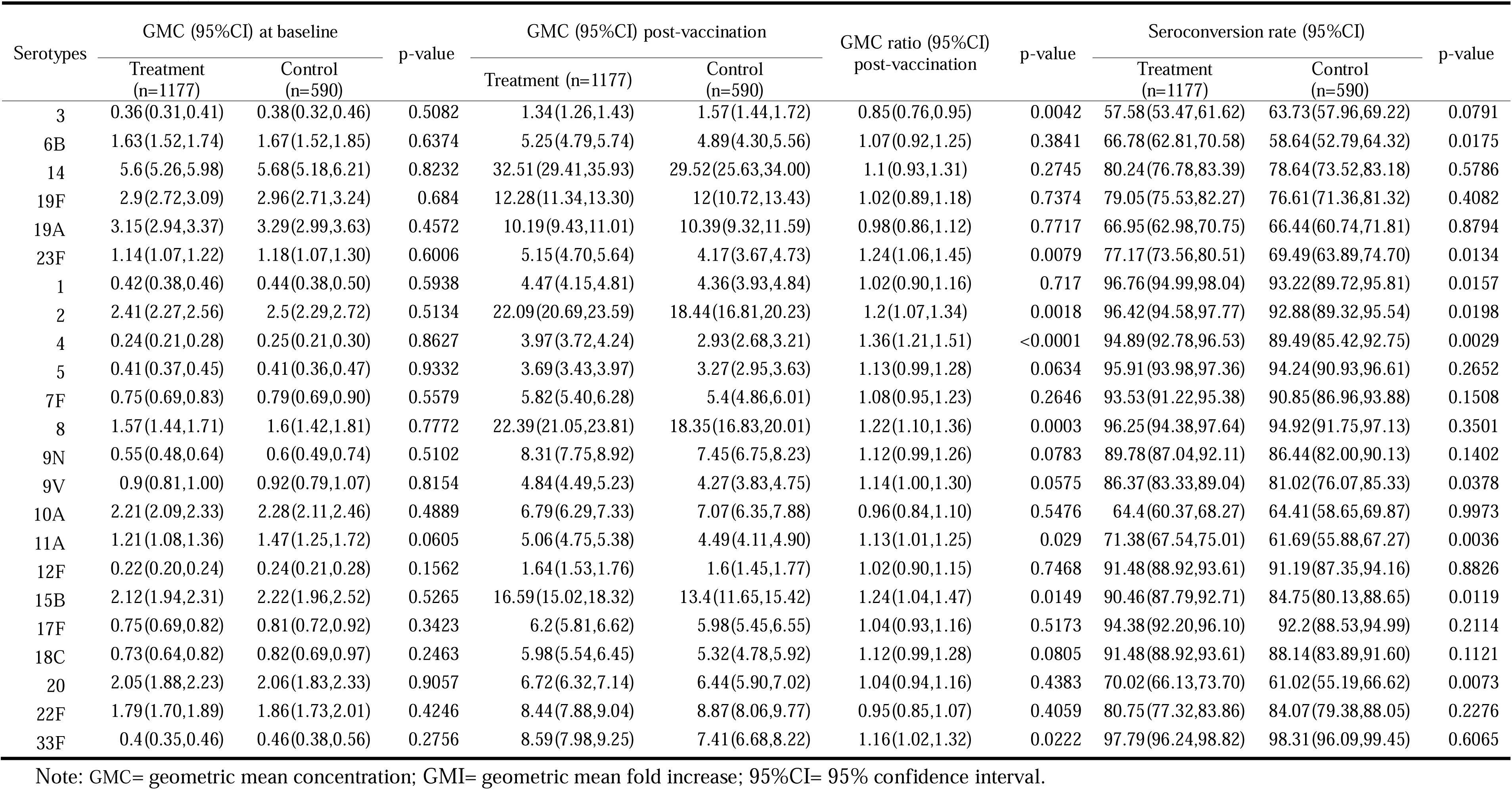
Serological Response of study participants aged 2-17 years to the 23-valent Pneumococcal Polysaccharide Vaccines.

**Supplementary Table 2.**
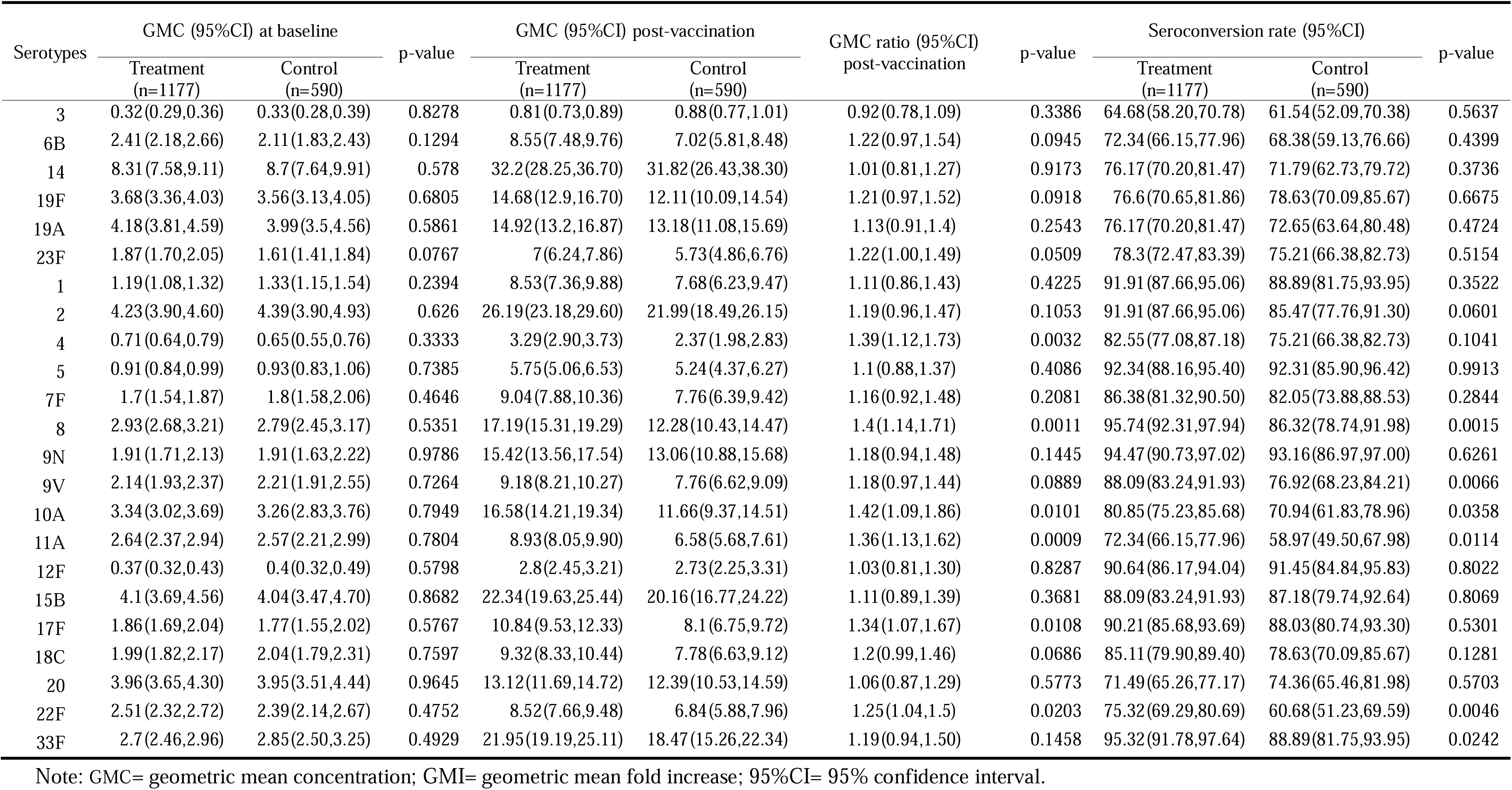
Serological Response of study participants aged 18-59 years to the 23-valent Pneumococcal Polysaccharide Vaccines.

**Supplementary Table 3.**
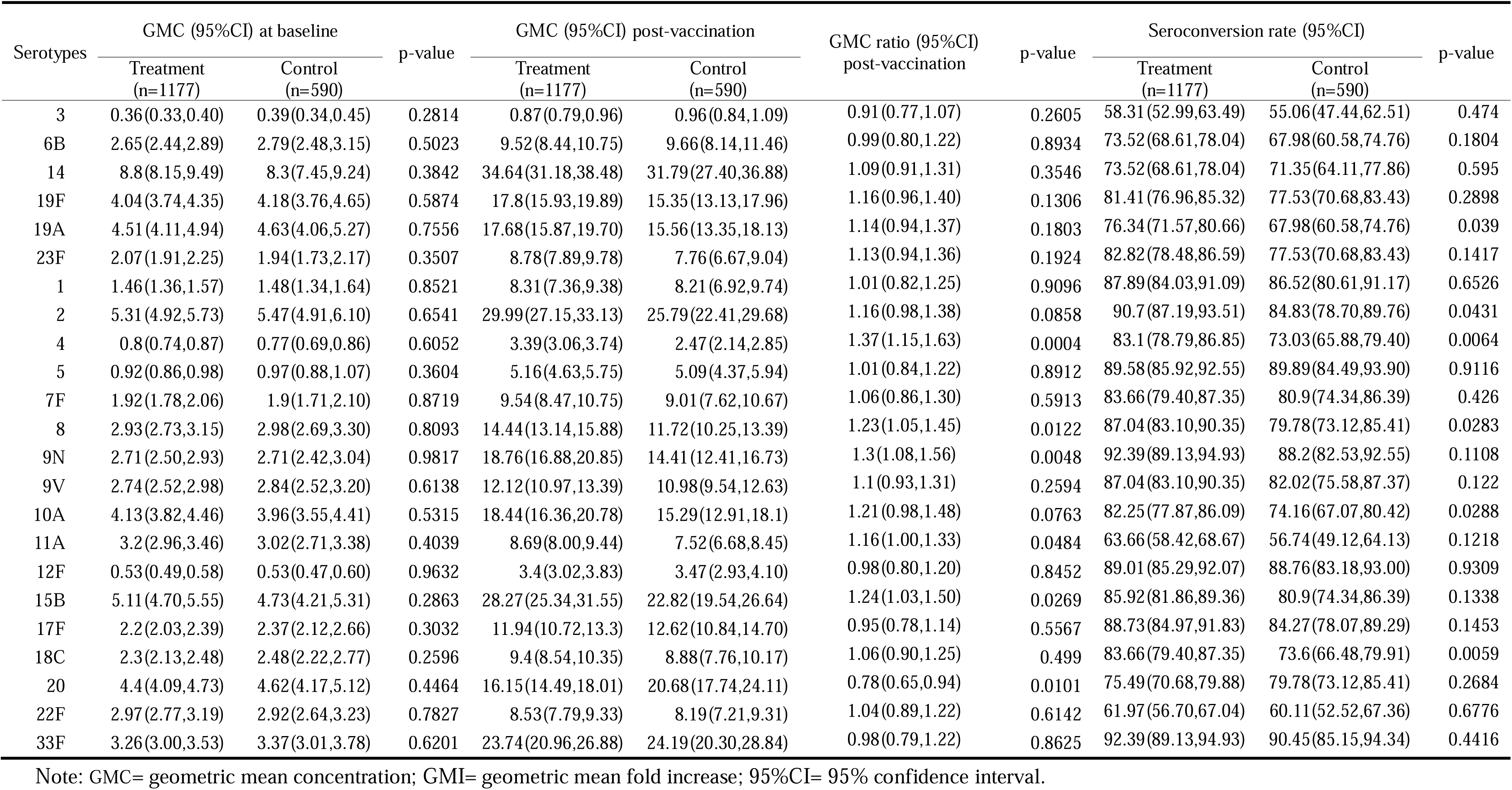
Serological Response of study participants aged 60+ years to the 23-valent Pneumococcal Polysaccharide Vaccines.

