## Supplementary Materials for "Safety and immunogenicity of 23-valent pneumococcal polysaccharide vaccine (PPV23) in Chinese children, adults and elderly: a phase 4, randomized, double-blind, active-controlled clinical trial"

This appendix has been provided by the authors to give readers additional information about their work.

### Supplementary tables

Supplementary Table 1. Serological Response of study participants aged 2-17 years to the 23-valent Pneumococcal Polysaccharide Vaccines

| Serotypes | GMC (95%CI) at baseline | | p-value | GMC (95%CI) post-vaccination | | GMC ratio (95%CI) post-vaccination | p-value | Seroconversion rate (95%CI) | | p-value |
| --- | --- | --- | --- | --- | --- | --- | --- | --- | --- | --- |
|  | Treatment (n=1177) | Control  (n=590) |  | Treatment (n=1177) | Control  (n=590) |  |  | Treatment (n=1177) | Control  (n=590) |  |
| 3 | 0.36(0.31,0.41) | 0.38(0.32,0.46) | 0.5082 | 1.34(1.26,1.43) | 1.57(1.44,1.72) | 0.85(0.76,0.95) | 0.0042 | 57.58(53.47,61.62) | 63.73(57.96,69.22) | 0.0791 |
| 6B | 1.63(1.52,1.74) | 1.67(1.52,1.85) | 0.6374 | 5.25(4.79,5.74) | 4.89(4.30,5.56) | 1.07(0.92,1.25) | 0.3841 | 66.78(62.81,70.58) | 58.64(52.79,64.32) | 0.0175 |
| 14 | 5.6(5.26,5.98) | 5.68(5.18,6.21) | 0.8232 | 32.51(29.41,35.93) | 29.52(25.63,34.00) | 1.1(0.93,1.31) | 0.2745 | 80.24(76.78,83.39) | 78.64(73.52,83.18) | 0.5786 |
| 19F | 2.9(2.72,3.09) | 2.96(2.71,3.24) | 0.684 | 12.28(11.34,13.30) | 12(10.72,13.43) | 1.02(0.89,1.18) | 0.7374 | 79.05(75.53,82.27) | 76.61(71.36,81.32) | 0.4082 |
| 19A | 3.15(2.94,3.37) | 3.29(2.99,3.63) | 0.4572 | 10.19(9.43,11.01) | 10.39(9.32,11.59) | 0.98(0.86,1.12) | 0.7717 | 66.95(62.98,70.75) | 66.44(60.74,71.81) | 0.8794 |
| 23F | 1.14(1.07,1.22) | 1.18(1.07,1.30) | 0.6006 | 5.15(4.70,5.64) | 4.17(3.67,4.73) | 1.24(1.06,1.45) | 0.0079 | 77.17(73.56,80.51) | 69.49(63.89,74.70) | 0.0134 |
| 1 | 0.42(0.38,0.46) | 0.44(0.38,0.50) | 0.5938 | 4.47(4.15,4.81) | 4.36(3.93,4.84) | 1.02(0.90,1.16) | 0.717 | 96.76(94.99,98.04) | 93.22(89.72,95.81) | 0.0157 |
| 2 | 2.41(2.27,2.56) | 2.5(2.29,2.72) | 0.5134 | 22.09(20.69,23.59) | 18.44(16.81,20.23) | 1.2(1.07,1.34) | 0.0018 | 96.42(94.58,97.77) | 92.88(89.32,95.54) | 0.0198 |
| 4 | 0.24(0.21,0.28) | 0.25(0.21,0.30) | 0.8627 | 3.97(3.72,4.24) | 2.93(2.68,3.21) | 1.36(1.21,1.51) | <0.0001 | 94.89(92.78,96.53) | 89.49(85.42,92.75) | 0.0029 |
| 5 | 0.41(0.37,0.45) | 0.41(0.36,0.47) | 0.9332 | 3.69(3.43,3.97) | 3.27(2.95,3.63) | 1.13(0.99,1.28) | 0.0634 | 95.91(93.98,97.36) | 94.24(90.93,96.61) | 0.2652 |
| 7F | 0.75(0.69,0.83) | 0.79(0.69,0.90) | 0.5579 | 5.82(5.40,6.28) | 5.4(4.86,6.01) | 1.08(0.95,1.23) | 0.2646 | 93.53(91.22,95.38) | 90.85(86.96,93.88) | 0.1508 |
| 8 | 1.57(1.44,1.71) | 1.6(1.42,1.81) | 0.7772 | 22.39(21.05,23.81) | 18.35(16.83,20.01) | 1.22(1.10,1.36) | 0.0003 | 96.25(94.38,97.64) | 94.92(91.75,97.13) | 0.3501 |
| 9N | 0.55(0.48,0.64) | 0.6(0.49,0.74) | 0.5102 | 8.31(7.75,8.92) | 7.45(6.75,8.23) | 1.12(0.99,1.26) | 0.0783 | 89.78(87.04,92.11) | 86.44(82.00,90.13) | 0.1402 |
| 9V | 0.9(0.81,1.00) | 0.92(0.79,1.07) | 0.8154 | 4.84(4.49,5.23) | 4.27(3.83,4.75) | 1.14(1.00,1.30) | 0.0575 | 86.37(83.33,89.04) | 81.02(76.07,85.33) | 0.0378 |
| 10A | 2.21(2.09,2.33) | 2.28(2.11,2.46) | 0.4889 | 6.79(6.29,7.33) | 7.07(6.35,7.88) | 0.96(0.84,1.10) | 0.5476 | 64.4(60.37,68.27) | 64.41(58.65,69.87) | 0.9973 |
| 11A | 1.21(1.08,1.36) | 1.47(1.25,1.72) | 0.0605 | 5.06(4.75,5.38) | 4.49(4.11,4.90) | 1.13(1.01,1.25) | 0.029 | 71.38(67.54,75.01) | 61.69(55.88,67.27) | 0.0036 |
| 12F | 0.22(0.20,0.24) | 0.24(0.21,0.28) | 0.1562 | 1.64(1.53,1.76) | 1.6(1.45,1.77) | 1.02(0.90,1.15) | 0.7468 | 91.48(88.92,93.61) | 91.19(87.35,94.16) | 0.8826 |
| 15B | 2.12(1.94,2.31) | 2.22(1.96,2.52) | 0.5265 | 16.59(15.02,18.32) | 13.4(11.65,15.42) | 1.24(1.04,1.47) | 0.0149 | 90.46(87.79,92.71) | 84.75(80.13,88.65) | 0.0119 |
| 17F | 0.75(0.69,0.82) | 0.81(0.72,0.92) | 0.3423 | 6.2(5.81,6.62) | 5.98(5.45,6.55) | 1.04(0.93,1.16) | 0.5173 | 94.38(92.20,96.10) | 92.2(88.53,94.99) | 0.2114 |
| 18C | 0.73(0.64,0.82) | 0.82(0.69,0.97) | 0.2463 | 5.98(5.54,6.45) | 5.32(4.78,5.92) | 1.12(0.99,1.28) | 0.0805 | 91.48(88.92,93.61) | 88.14(83.89,91.60) | 0.1121 |
| 20 | 2.05(1.88,2.23) | 2.06(1.83,2.33) | 0.9057 | 6.72(6.32,7.14) | 6.44(5.90,7.02) | 1.04(0.94,1.16) | 0.4383 | 70.02(66.13,73.70) | 61.02(55.19,66.62) | 0.0073 |
| 22F | 1.79(1.70,1.89) | 1.86(1.73,2.01) | 0.4246 | 8.44(7.88,9.04) | 8.87(8.06,9.77) | 0.95(0.85,1.07) | 0.4059 | 80.75(77.32,83.86) | 84.07(79.38,88.05) | 0.2276 |
| 33F | 0.4(0.35,0.46) | 0.46(0.38,0.56) | 0.2756 | 8.59(7.98,9.25) | 7.41(6.68,8.22) | 1.16(1.02,1.32) | 0.0222 | 97.79(96.24,98.82) | 98.31(96.09,99.45) | 0.6065 |

Note: GMC= geometric mean concentration; GMI= geometric mean fold increase; 95%CI= 95% confidence interval.

Supplementary Table 2. Serological Response of study participants aged 18-59 years to the 23-valent Pneumococcal Polysaccharide Vaccines

| Serotypes | GMC (95%CI) at baseline | | p-value | GMC (95%CI) post-vaccination | | GMC ratio (95%CI) post-vaccination | p-value | Seroconversion rate (95%CI) | | p-value |
| --- | --- | --- | --- | --- | --- | --- | --- | --- | --- | --- |
|  | Treatment (n=1177) | Control  (n=590) |  | Treatment (n=1177) | Control  (n=590) |  |  | Treatment (n=1177) | Control  (n=590) |  |
| 3 | 0.32(0.29,0.36) | 0.33(0.28,0.39) | 0.8278 | 0.81(0.73,0.89) | 0.88(0.77,1.01) | 0.92(0.78,1.09) | 0.3386 | 64.68(58.20,70.78) | 61.54(52.09,70.38) | 0.5637 |
| 6B | 2.41(2.18,2.66) | 2.11(1.83,2.43) | 0.1294 | 8.55(7.48,9.76) | 7.02(5.81,8.48) | 1.22(0.97,1.54) | 0.0945 | 72.34(66.15,77.96) | 68.38(59.13,76.66) | 0.4399 |
| 14 | 8.31(7.58,9.11) | 8.7(7.64,9.91) | 0.578 | 32.2(28.25,36.70) | 31.82(26.43,38.30) | 1.01(0.81,1.27) | 0.9173 | 76.17(70.20,81.47) | 71.79(62.73,79.72) | 0.3736 |
| 19F | 3.68(3.36,4.03) | 3.56(3.13,4.05) | 0.6805 | 14.68(12.9,16.70) | 12.11(10.09,14.54) | 1.21(0.97,1.52) | 0.0918 | 76.6(70.65,81.86) | 78.63(70.09,85.67) | 0.6675 |
| 19A | 4.18(3.81,4.59) | 3.99(3.5,4.56) | 0.5861 | 14.92(13.2,16.87) | 13.18(11.08,15.69) | 1.13(0.91,1.4) | 0.2543 | 76.17(70.20,81.47) | 72.65(63.64,80.48) | 0.4724 |
| 23F | 1.87(1.70,2.05) | 1.61(1.41,1.84) | 0.0767 | 7(6.24,7.86) | 5.73(4.86,6.76) | 1.22(1.00,1.49) | 0.0509 | 78.3(72.47,83.39) | 75.21(66.38,82.73) | 0.5154 |
| 1 | 1.19(1.08,1.32) | 1.33(1.15,1.54) | 0.2394 | 8.53(7.36,9.88) | 7.68(6.23,9.47) | 1.11(0.86,1.43) | 0.4225 | 91.91(87.66,95.06) | 88.89(81.75,93.95) | 0.3522 |
| 2 | 4.23(3.90,4.60) | 4.39(3.90,4.93) | 0.626 | 26.19(23.18,29.60) | 21.99(18.49,26.15) | 1.19(0.96,1.47) | 0.1053 | 91.91(87.66,95.06) | 85.47(77.76,91.30) | 0.0601 |
| 4 | 0.71(0.64,0.79) | 0.65(0.55,0.76) | 0.3333 | 3.29(2.90,3.73) | 2.37(1.98,2.83) | 1.39(1.12,1.73) | 0.0032 | 82.55(77.08,87.18) | 75.21(66.38,82.73) | 0.1041 |
| 5 | 0.91(0.84,0.99) | 0.93(0.83,1.06) | 0.7385 | 5.75(5.06,6.53) | 5.24(4.37,6.27) | 1.1(0.88,1.37) | 0.4086 | 92.34(88.16,95.40) | 92.31(85.90,96.42) | 0.9913 |
| 7F | 1.7(1.54,1.87) | 1.8(1.58,2.06) | 0.4646 | 9.04(7.88,10.36) | 7.76(6.39,9.42) | 1.16(0.92,1.48) | 0.2081 | 86.38(81.32,90.50) | 82.05(73.88,88.53) | 0.2844 |
| 8 | 2.93(2.68,3.21) | 2.79(2.45,3.17) | 0.5351 | 17.19(15.31,19.29) | 12.28(10.43,14.47) | 1.4(1.14,1.71) | 0.0011 | 95.74(92.31,97.94) | 86.32(78.74,91.98) | 0.0015 |
| 9N | 1.91(1.71,2.13) | 1.91(1.63,2.22) | 0.9786 | 15.42(13.56,17.54) | 13.06(10.88,15.68) | 1.18(0.94,1.48) | 0.1445 | 94.47(90.73,97.02) | 93.16(86.97,97.00) | 0.6261 |
| 9V | 2.14(1.93,2.37) | 2.21(1.91,2.55) | 0.7264 | 9.18(8.21,10.27) | 7.76(6.62,9.09) | 1.18(0.97,1.44) | 0.0889 | 88.09(83.24,91.93) | 76.92(68.23,84.21) | 0.0066 |
| 10A | 3.34(3.02,3.69) | 3.26(2.83,3.76) | 0.7949 | 16.58(14.21,19.34) | 11.66(9.37,14.51) | 1.42(1.09,1.86) | 0.0101 | 80.85(75.23,85.68) | 70.94(61.83,78.96) | 0.0358 |
| 11A | 2.64(2.37,2.94) | 2.57(2.21,2.99) | 0.7804 | 8.93(8.05,9.90) | 6.58(5.68,7.61) | 1.36(1.13,1.62) | 0.0009 | 72.34(66.15,77.96) | 58.97(49.50,67.98) | 0.0114 |
| 12F | 0.37(0.32,0.43) | 0.4(0.32,0.49) | 0.5798 | 2.8(2.45,3.21) | 2.73(2.25,3.31) | 1.03(0.81,1.30) | 0.8287 | 90.64(86.17,94.04) | 91.45(84.84,95.83) | 0.8022 |
| 15B | 4.1(3.69,4.56) | 4.04(3.47,4.70) | 0.8682 | 22.34(19.63,25.44) | 20.16(16.77,24.22) | 1.11(0.89,1.39) | 0.3681 | 88.09(83.24,91.93) | 87.18(79.74,92.64) | 0.8069 |
| 17F | 1.86(1.69,2.04) | 1.77(1.55,2.02) | 0.5767 | 10.84(9.53,12.33) | 8.1(6.75,9.72) | 1.34(1.07,1.67) | 0.0108 | 90.21(85.68,93.69) | 88.03(80.74,93.30) | 0.5301 |
| 18C | 1.99(1.82,2.17) | 2.04(1.79,2.31) | 0.7597 | 9.32(8.33,10.44) | 7.78(6.63,9.12) | 1.2(0.99,1.46) | 0.0686 | 85.11(79.90,89.40) | 78.63(70.09,85.67) | 0.1281 |
| 20 | 3.96(3.65,4.30) | 3.95(3.51,4.44) | 0.9645 | 13.12(11.69,14.72) | 12.39(10.53,14.59) | 1.06(0.87,1.29) | 0.5773 | 71.49(65.26,77.17) | 74.36(65.46,81.98) | 0.5703 |
| 22F | 2.51(2.32,2.72) | 2.39(2.14,2.67) | 0.4752 | 8.52(7.66,9.48) | 6.84(5.88,7.96) | 1.25(1.04,1.5) | 0.0203 | 75.32(69.29,80.69) | 60.68(51.23,69.59) | 0.0046 |
| 33F | 2.7(2.46,2.96) | 2.85(2.50,3.25) | 0.4929 | 21.95(19.19,25.11) | 18.47(15.26,22.34) | 1.19(0.94,1.50) | 0.1458 | 95.32(91.78,97.64) | 88.89(81.75,93.95) | 0.0242 |

Note: GMC= geometric mean concentration; GMI= geometric mean fold increase; 95%CI= 95% confidence interval.

Supplementary Table 3. Serological Response of study participants aged 60+ years to the 23-valent Pneumococcal Polysaccharide Vaccines

| Serotypes | GMC (95%CI) at baseline | | p-value | GMC (95%CI) post-vaccination | | GMC ratio (95%CI) post-vaccination | p-value | Seroconversion rate (95%CI) | | p-value |
| --- | --- | --- | --- | --- | --- | --- | --- | --- | --- | --- |
|  | Treatment (n=1177) | Control  (n=590) |  | Treatment (n=1177) | Control  (n=590) |  |  | Treatment (n=1177) | Control  (n=590) |  |
| 3 | 0.36(0.33,0.40) | 0.39(0.34,0.45) | 0.2814 | 0.87(0.79,0.96) | 0.96(0.84,1.09) | 0.91(0.77,1.07) | 0.2605 | 58.31(52.99,63.49) | 55.06(47.44,62.51) | 0.474 |
| 6B | 2.65(2.44,2.89) | 2.79(2.48,3.15) | 0.5023 | 9.52(8.44,10.75) | 9.66(8.14,11.46) | 0.99(0.80,1.22) | 0.8934 | 73.52(68.61,78.04) | 67.98(60.58,74.76) | 0.1804 |
| 14 | 8.8(8.15,9.49) | 8.3(7.45,9.24) | 0.3842 | 34.64(31.18,38.48) | 31.79(27.40,36.88) | 1.09(0.91,1.31) | 0.3546 | 73.52(68.61,78.04) | 71.35(64.11,77.86) | 0.595 |
| 19F | 4.04(3.74,4.35) | 4.18(3.76,4.65) | 0.5874 | 17.8(15.93,19.89) | 15.35(13.13,17.96) | 1.16(0.96,1.40) | 0.1306 | 81.41(76.96,85.32) | 77.53(70.68,83.43) | 0.2898 |
| 19A | 4.51(4.11,4.94) | 4.63(4.06,5.27) | 0.7556 | 17.68(15.87,19.70) | 15.56(13.35,18.13) | 1.14(0.94,1.37) | 0.1803 | 76.34(71.57,80.66) | 67.98(60.58,74.76) | 0.039 |
| 23F | 2.07(1.91,2.25) | 1.94(1.73,2.17) | 0.3507 | 8.78(7.89,9.78) | 7.76(6.67,9.04) | 1.13(0.94,1.36) | 0.1924 | 82.82(78.48,86.59) | 77.53(70.68,83.43) | 0.1417 |
| 1 | 1.46(1.36,1.57) | 1.48(1.34,1.64) | 0.8521 | 8.31(7.36,9.38) | 8.21(6.92,9.74) | 1.01(0.82,1.25) | 0.9096 | 87.89(84.03,91.09) | 86.52(80.61,91.17) | 0.6526 |
| 2 | 5.31(4.92,5.73) | 5.47(4.91,6.10) | 0.6541 | 29.99(27.15,33.13) | 25.79(22.41,29.68) | 1.16(0.98,1.38) | 0.0858 | 90.7(87.19,93.51) | 84.83(78.70,89.76) | 0.0431 |
| 4 | 0.8(0.74,0.87) | 0.77(0.69,0.86) | 0.6052 | 3.39(3.06,3.74) | 2.47(2.14,2.85) | 1.37(1.15,1.63) | 0.0004 | 83.1(78.79,86.85) | 73.03(65.88,79.40) | 0.0064 |
| 5 | 0.92(0.86,0.98) | 0.97(0.88,1.07) | 0.3604 | 5.16(4.63,5.75) | 5.09(4.37,5.94) | 1.01(0.84,1.22) | 0.8912 | 89.58(85.92,92.55) | 89.89(84.49,93.90) | 0.9116 |
| 7F | 1.92(1.78,2.06) | 1.9(1.71,2.10) | 0.8719 | 9.54(8.47,10.75) | 9.01(7.62,10.67) | 1.06(0.86,1.30) | 0.5913 | 83.66(79.40,87.35) | 80.9(74.34,86.39) | 0.426 |
| 8 | 2.93(2.73,3.15) | 2.98(2.69,3.30) | 0.8093 | 14.44(13.14,15.88) | 11.72(10.25,13.39) | 1.23(1.05,1.45) | 0.0122 | 87.04(83.10,90.35) | 79.78(73.12,85.41) | 0.0283 |
| 9N | 2.71(2.50,2.93) | 2.71(2.42,3.04) | 0.9817 | 18.76(16.88,20.85) | 14.41(12.41,16.73) | 1.3(1.08,1.56) | 0.0048 | 92.39(89.13,94.93) | 88.2(82.53,92.55) | 0.1108 |
| 9V | 2.74(2.52,2.98) | 2.84(2.52,3.20) | 0.6138 | 12.12(10.97,13.39) | 10.98(9.54,12.63) | 1.1(0.93,1.31) | 0.2594 | 87.04(83.10,90.35) | 82.02(75.58,87.37) | 0.122 |
| 10A | 4.13(3.82,4.46) | 3.96(3.55,4.41) | 0.5315 | 18.44(16.36,20.78) | 15.29(12.91,18.1) | 1.21(0.98,1.48) | 0.0763 | 82.25(77.87,86.09) | 74.16(67.07,80.42) | 0.0288 |
| 11A | 3.2(2.96,3.46) | 3.02(2.71,3.38) | 0.4039 | 8.69(8.00,9.44) | 7.52(6.68,8.45) | 1.16(1.00,1.33) | 0.0484 | 63.66(58.42,68.67) | 56.74(49.12,64.13) | 0.1218 |
| 12F | 0.53(0.49,0.58) | 0.53(0.47,0.60) | 0.9632 | 3.4(3.02,3.83) | 3.47(2.93,4.10) | 0.98(0.80,1.20) | 0.8452 | 89.01(85.29,92.07) | 88.76(83.18,93.00) | 0.9309 |
| 15B | 5.11(4.70,5.55) | 4.73(4.21,5.31) | 0.2863 | 28.27(25.34,31.55) | 22.82(19.54,26.64) | 1.24(1.03,1.50) | 0.0269 | 85.92(81.86,89.36) | 80.9(74.34,86.39) | 0.1338 |
| 17F | 2.2(2.03,2.39) | 2.37(2.12,2.66) | 0.3032 | 11.94(10.72,13.3) | 12.62(10.84,14.70) | 0.95(0.78,1.14) | 0.5567 | 88.73(84.97,91.83) | 84.27(78.07,89.29) | 0.1453 |
| 18C | 2.3(2.13,2.48) | 2.48(2.22,2.77) | 0.2596 | 9.4(8.54,10.35) | 8.88(7.76,10.17) | 1.06(0.90,1.25) | 0.499 | 83.66(79.40,87.35) | 73.6(66.48,79.91) | 0.0059 |
| 20 | 4.4(4.09,4.73) | 4.62(4.17,5.12) | 0.4464 | 16.15(14.49,18.01) | 20.68(17.74,24.11) | 0.78(0.65,0.94) | 0.0101 | 75.49(70.68,79.88) | 79.78(73.12,85.41) | 0.2684 |
| 22F | 2.97(2.77,3.19) | 2.92(2.64,3.23) | 0.7827 | 8.53(7.79,9.33) | 8.19(7.21,9.31) | 1.04(0.89,1.22) | 0.6142 | 61.97(56.70,67.04) | 60.11(52.52,67.36) | 0.6776 |
| 33F | 3.26(3.00,3.53) | 3.37(3.01,3.78) | 0.6201 | 23.74(20.96,26.88) | 24.19(20.30,28.84) | 0.98(0.79,1.22) | 0.8625 | 92.39(89.13,94.93) | 90.45(85.15,94.34) | 0.4416 |

Note: GMC= geometric mean concentration; GMI= geometric mean fold increase; 95%CI= 95% confidence interval.
